# CRISPR-Cas12a2-based rapid and sensitive detection system for target nucleic acid

**DOI:** 10.1101/2024.09.20.24314102

**Authors:** Helin Yu, Meng Feng, Chuncao Liu, Feifei Wang, Guodong Sui, Wenwen Jing, Xunjia Cheng

**Affiliations:** Shanghai Institute of Infectious Disease and Biosecurity, Fudan University, Shanghai 200032, China; Department of Medical Microbiology and Parasitology, School of Basic Medical Sciences, Fudan University, Shanghai 20032, China; Department of Environmental Science and Engineering, Fudan University, Shanghai 200438, China

**Keywords:** CRISPR-SuCas12a2, nucleic acid detection, ISOthermal reaction, target RNA, fluorescence quantitation

## Abstract

Infectious diseases are extremely important public health issues, where the design of effective, rapid, and convenient detection platforms is critical. In this study, we used conventional PCR coupled with SuCas12a2, a novel Cas12 family RNA-targeting nuclease, to develop a detection approach. SuCas12a2 possesses collateral cleavage activity and cuts the additional single-stranded RNA (ssRNA) added to the reaction system once the ternary complex RNA-SuCas12a2-CRISPR RNA (crRNA) is formed. SuCas12a2 is specifically activated, where the cleaved fluorescent-labeled probes release fluorescent signals, with the strength of the fluorescent signal being proportional to the concentration of nucleic acids specifically bound to crRNA. Simultaneous transcription and SuCas12a2 detection can be performed in a single tube by introducing the T7 promoter sequence into the forward primer. *Entamoeba histolytica* was used to evaluate the performance of the platform. PCR-SuCas12a2 has excellent capabilities, including high specificity with no cross-reactivity from other species and ultra-sensitivity that achieves a detection of one copy per reaction. There were five samples from amoebiasis patients confirmed by indirect immunofluorescence assay that were used as proof specimens, where the PCR-SuCas12a2 assay demonstrated 100% specificity. Furthermore, we replaced conventional PCR with recombinase polymerase amplification (RPA) to simplify the procedure for producing amplicons harboring the T7 promoter sequence. The sensitivity of the RPA-SuCas12a2 assay was 10^2^ copies per reaction, which was inferior to PCR-SuCas12a2, and demonstrated 100% specificity. The technique shows robust performance and suggests great potential for point-of-care testing of other pathogens to facilitate effective management and control of the spread of diseases.

Figure abstract

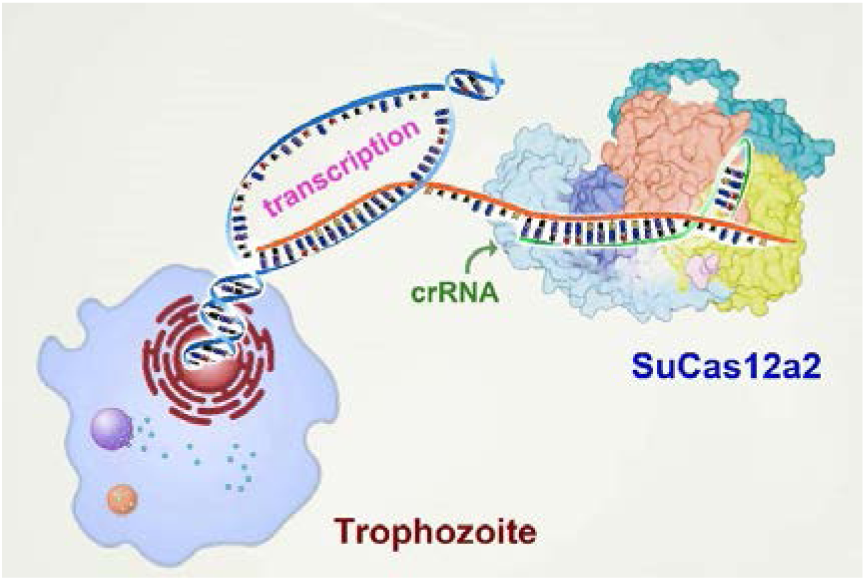

## 1. Introduction

Infectious diseases have always been a significant challenge to global public health, with significant and rapid transmission posing a serious threat to human health and socioeconomic development [1]. Immediate and accurate diagnosis is crucial for the effective prevention and control of infectious diseases [2]. With the rapid development of biotechnology, high-throughput, rapid, and specific diagnostic methods have emerged, bringing revolutionary changes to the diagnosis of infectious diseases [3,4]. High-throughput diagnostic methods are usually based on large-scale parallel detection technology that can simultaneously detect multiple samples or multiple pathogen targets [5–7]. For example, gene-chip technology can integrate thousands of nucleic acid probes on a single chip to simultaneously detect multiple pathogenic genes [8]. Therefore, high-throughput, rapid, and specific diagnostic methods are essential to diagnose infectious diseases, not only improving the efficiency and accuracy of diagnosis but also effectively monitoring and controlling the spread and prevalence of infectious diseases.

Serological methods for the diagnosis of pathogens are based on antigen-antibody reactions, such as enzyme-linked immunosorbent assay (ELISA), which are feasible only after the antibodies have been produced and do not determine whether they are currently in a state of infection [2,9,10]. Nucleic acid detection, a crucial tool in modern medical diagnostics, plays a pivotal role in pathogen detection, disease diagnosis, and genetic analysis, owing to its high specificity and sensitivity. Methodologies have been continuously enriched and refined from simple nucleic acid hybridization to high-throughput sequencing with precise quantitative PCR. However, nucleic acid detection faces several challenges. Nucleic acid detection has become a crucial diagnostic tool in preventing and controlling infectious diseases, such as acquired immune deficiency syndrome, hepatitis B, severe acute respiratory syndrome coronavirus 2 (SARS-CoV-2), and bacterial infections [11–13].

In recent years, the emergence of CRISPR-Cas technology has provided new opportunities to diagnose infectious diseases. CRISPR-Cas enzymes have significant advantages in the diagnosis of infectious diseases, are capable of detecting pathogen nucleic acids at extremely low concentrations, and greatly enhance diagnostic accuracy and sensitivity. For example, to detect certain viral infections, traditional methods may require a large number of viral particles, whereas CRISPR-Cas technology can detect nucleic acids in a single viral particle. By precisely designing guide RNAs, specific gene sequences of pathogens can be recognized, and cross-reactions with other similar pathogens or host genes can be avoided, thus achieving high specificity. The detection process is relatively simple and rapid, and CRISPR-Cas diagnostics are typically completed within hours, meaning relatively high speed and convenience. By designing multiple guide RNAs, the simultaneous detection of multiple pathogens can be achieved, thereby improving diagnostic efficiency. DNA Endonuclease-Targeted CRISPR Trans Reporter (DETECTR) and Specific High-sensitivity Enzymatic Reporter UnLOCKing (SHERLOCK) are the classical detection tools based on Cas12 and Cas13, respectively [14–17]. They are widely used to detect various pathogens such as SARS-CoV-2 [18], Zika virus (ZIKV) [19,20], Dengue virus [21,22], and Human immunodeficiency virus [23]. Generally, these approaches are coupled with isothermal amplification approaches, such as recombinase polymerase amplification (RPA) [24], rolling circle amplification (RCA) [25], and loop-mediated isothermal amplification (LAMP) [26,27], to amplify target DNA fragments with a few copies of the template substrate to the limit of detection (LoD) in a short time at a single temperature.

Novel Cas effector proteins have gradually been identified, and new detection tools have been exploited with the development of advanced molecular techniques [28]. SuCas12a2, with a molecular mass of 145.6 kDa, has recently been discovered and belongs to the Cas12 family. Similar to Cas12a and Cas12b, Cas12a2 has double-stranded DNA (dsDNA)-specific activity and non-specific collateral cleavage activity. SuCas12a2, however, is an RNA-guided single nuclease, rather than dsDNA-guided, and effectively degrades dsDNA, single-stranded DNA (ssDNA), and ssRNA once SuCas12a2 recognizes the protospacer-flanking sequence (PFS, 5’-GAAAG-3’), which can be used to provide alternative diagnostic tools [29,30]. New CRISPR-Cas enzymes are continuously being discovered, and research on diagnostic technologies is rapidly advancing. New detection signal output methods, such as fluorescence, colorimetry, and electrochemistry, are being continuously developed to enhance the convenience and visualization of detection and achieve innovative detection methods. Moreover, multimodal forms have been developed in combination with other technologies.

This work describes our design of a sensitive, specific, rapid, and convenient pathogen detection system utilizing SuCas12a2. *Entamoeba histolytica* (*E. histolytica*) was used as a model pathogen to validate the assay. PCR was performed to amplify the *E. histolytica* peroxiredoxin gene (*Eh*Prx) using primers harboring the T7 promoter sequence, which was screened with SuCas12a2 combined with crRNA for detection. Furthermore, we replaced conventional PCR with RPA to simplify the procedure by coupling RPA and the CRISPR-SuCas12a2 system.

This study aimed to demonstrate the high performance of our assay, as this new molecular scissor tool demonstrates the potential of novel CRISPR-Cas-based diagnostics for pathogens with varying selectivity.

## 2. Materials and methods

### 2.1 Ethical considerations

The serum samples, feces samples, and liver tissues of five patients were collected from hospitals affiliated to Fudan University. In accordance with the Helsinki Declaration, the study protocol was subject to the approval of the Medical Ethics Committee of School of Basic Medical Science, Fudan University (permit number: 2018Y022). All participants were recruited with written informed consent prior to the study.

### 2.2 Expression and purification of SuCas12a2

The pET52b-SuCas12a2 plasmids were transformed into *E. coli* Nico21 (DE3) competent cells and clones harboring SuCas12a2 plasmids were isolated from the plates in LB broth. A final concentration of 1 mM Isopropylthio-*β*-galactoside (IPTG) was supplemented to the LB medium, and protein expression was induced for 18 hours at 18℃.

The protein was purified by affinity chromatography and molecular exclusion chromatography according to the manufacturer’s instructions. The purified SuCas12a2 was concentrated utilizing a 100 kDa MWKO vivaspin 500 concentrator, and molecular-exclusion chromatography was performed using a HiLoad^TM^ 16/600 Superdex^TM^ 200 pg column for further purification. The elution buffer was exchanged for a storage buffer (100 mM HEPES pH 7.2, 150 mM KCl, 2mM MgCl_2_, 10% glycerol, 1 mM DTT), and the protein was flash freezed and stored at -80℃ until further use. SDS-PAGE and coomassie blue staining were used to identify the purity of the protein, while the Qubit protein assay kit (Thermo Fisher Scientific; Q33211) was utilized for protein quantification.

### 2.3 Plasmid cleavage assay

A plasmid cleavage assay was performed using the pUC19. A total of 10 μl reaction was used, containing 50 nM, 20 nM, 10 nM of SuCas12a2 with corresponding concentrations of crRNA, target RNA, and 4 nM supercoiled pUC19 plasmid mixed in 1 × HOLMES buffer 1 (TOLOBIO; 32005). The mixture was incubated for different time points at 1, 2, 5, 10, 15, 30, and 60 min at 37℃ and then heated at 75℃ for 5 min to inactivate the SuCas12a2. 0.8% nucleic acid agarose gel was used to visualize plasmid configuration.

### 2.4 Target ds-DNA template preparation and pre-PCR reaction

All target dsDNA oligo fragments (*prx*) containing the T7 promoter were amplified using the Ex Taq enzyme (TAKARA; RR001). Clinical samples’ or trophozoites’ genomic DNA (gDNA) was extracted using a DNeasy Blood & Tissue Kit (Qiagen; 69506). Nuclease-free water was used as a negative control. Primers were designed using Primer Premier 5, and the T7 promoter was introduced into the target dsDNA fragment using forward primer. In addition, the amplicon contained a sequence that combines with crRNA. A total of 20 μl PCR reaction volume included 2 μl of 10 × Ex Taq buffer, 1.6 μl of dNTP mixture (2.5 mM each), 2 μl of 10 μM *prx-*dsDNA forward and reverse primers, 50 ng of substrate gDNA, and 0.1 μl of Taq DNA polymerase, with nuclease-free water added to reach a total volume of 20 μl. PCR was performed as the following procedure: 94℃ 3 min, and 40 cycles for 94℃ 15 s, 58℃ 30 s, 72℃ 30 s, and 72℃ for 7 min. The amplified product was purified using the AxyPrep^TM^ PCR Clean-up Kit (Axygen; 34617KB1) according to the manufacturer’s instructions. The purity of target dsDNA after clean-up was identified by 2% nucleic acid agarose gel and quantified by Qubit dsDNA HS Assay Kit (Thermo Fisher Scientific; Q32850). To limit the detection assay, the substrate gDNA was diluted in multiple ratios to different copies per microliter.

### 2.5 RPA reaction

Primers containing the T7 promoter for RPA were designed using Primer Premier 5 and the most suitable primers were selected according to the instructions of the TwistAmp Basic (TwistDx; 844803) primer screening section. The amplification substrate was replaced with *E. histolytica* gDNA by pET19b-*Eh*prx plasmid, and different concentrations of 1 μl plasmid were utilized in the LoD assay. RPA amplification of clinical samples was performed using 2 μl of the template. All reactions were incubated in a water bath of 37℃ for 40 min, and purification of the RPA products was unnecessary.

### 2.6 CRISPR-SuCas12a2 reaction for fluorescence detection

The SuCas12a2 *trans*-cleavage assay was performed after amplicons were obtained. A total of 20 μl reaction mixture was created containing 2 μl of 10 × HOLMES buffer 1, 50 nM purified SuCas12a2, 200 nM crRNA1-*prx* (GENCEFE Biotech), 1 mM NTP mixture (Sangon Biotech; B600057), 2,000 U/mL recombinant RNase inhibitor (TaKaRa; 2313B), 5,000 U/mL T7 RNA polymerase (TaKaRa; 2540B), 1 μl of PCR product, and 250 nM ssRNA reporter (5’/6-FAM-UUUUU-BHQ1/3’) synthesized by GENCEFE Biotech, and the remaining volume with nuclease-free water. The mixture was transferred into a Semi-skirt PCR plate (VIOX Scientific; VP2021-C) and incubated at 37℃ in the ABI 7500 instrument (Thermo Fisher Scientific) for 45 min, during which the fluorescence intensity was measured every 30 seconds. Three technical replicates were used for each assay.

### 2.7 Indirect immunofluorescence assay for clinical samples

An indirect immunofluorescence assay (IFA) was performed using five clinical samples as positive controls. A total of 5×10^3^ fixed trophozoites were smeared onto cytoslides using a Cytospin 4 cytocentrifuge (Thermo Fisher Scientific, Waltham, MA, USA) at 800 rpm for 10 minutes and then blocked with 5% bovine serum albumin (BSA) for 15 min. The serum of suspected infected *E. histolytica* was diluted with 5% BSA 1:64, and the serum of healthy individuals was used as a negative control. Serum samples were added to each slide and incubated at room temperature for 30 min. Alexa Fluor 488 anti-human IgG H&L (1:100, Thermo Fisher Scientific; A18805) was used as the secondary antibody. Cytoslides were observed under an inverted microscope (Olympus).

## 3. Results

### 3.1 Recombinant protein SuCas12a2 purification and *trans*-cleavage activity

The SuCas12a2 sequence was synthesized and inserted into a pET52b (+) expression vector after NotI/BamHI digestion. The SuCas12a2 protein, containing C-terminal 10×His tag with a molecular weight of 145.6 kDa, was abundantly expressed in *Escherichia coli* Nico21 (DE3) and easily detectable in the supernatant and purified by Ni-NTA and molecular exclusion chromatography (Figure 1a). Then the monomeric SuCas12a2 protein was collected and concentrated (Figure 1b). To assess the dsDNA cleavage capacity of the recombinant SuCas12a2, we conducted pUC19 plasmid cleavage experiments to challenge with SuCas12a2 at various concentrations and incubation times (Figure 1c). Partial nicking and linearization of the supercoiled pUC19 plasmid were conducted within one minute with SuCas12a2 present at a concentration of 10 nM. Complete degradation of the plasmid occurred after 60 min at 50 nM, 20 nM, and 10 nM concentrations tested. Furthermore, increasing the concentration of SuCas12a2 or prolonging the incubation duration significantly enhanced plasmid degradation (Figure 1d). These findings demonstrated that SuCas12a2-mediated dsDNA cleavage is both time- and concentration-dependent. Overall, these data suggested that the purified SuCas12a2 protein was functionally stable and could be used to develop novel diagnostic platforms.

**Figure 1.**
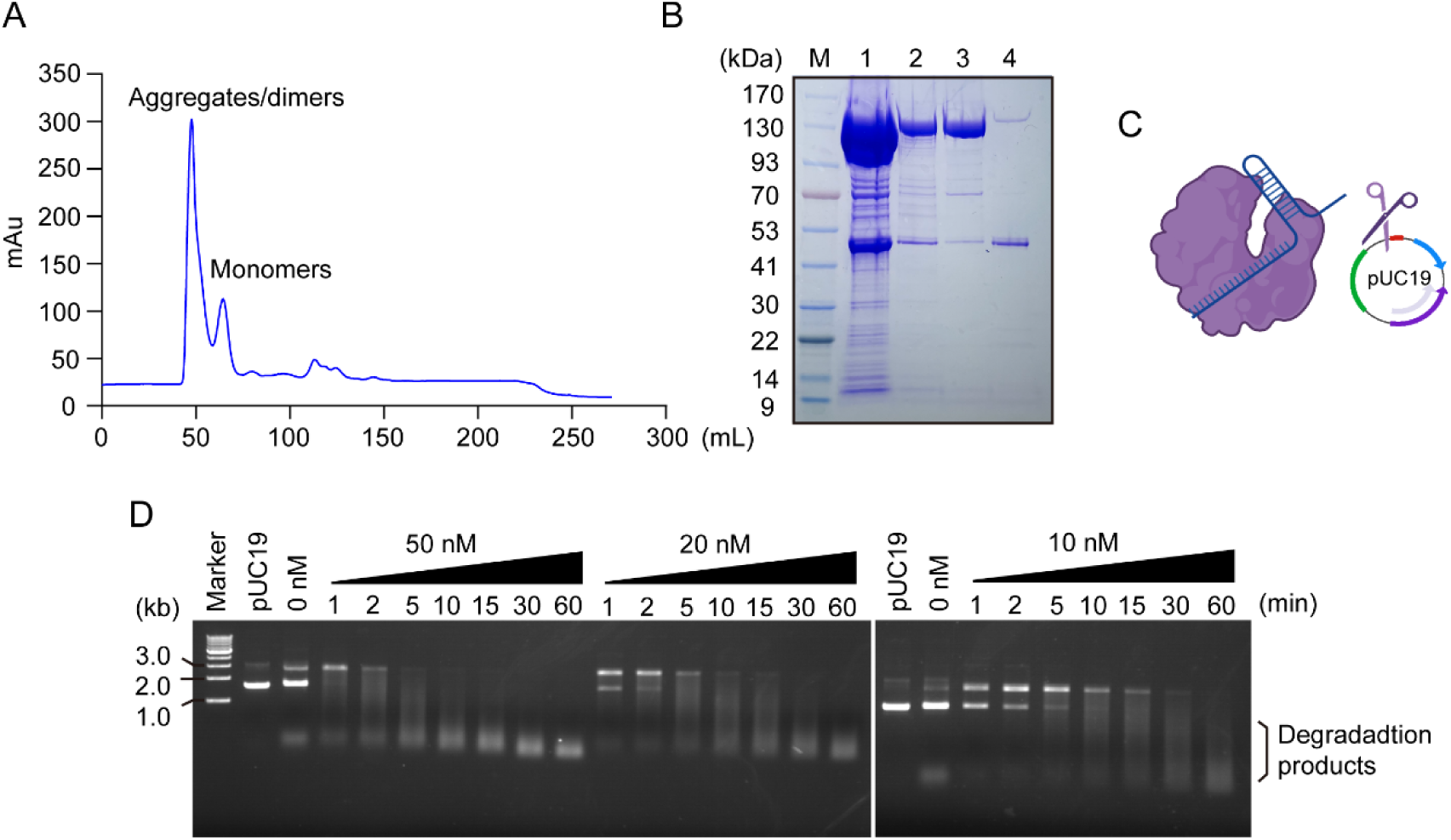
Purification and non-specific cleavage activity verification of SuCas12a2 protein. (A). Molecular exclusion chromatography of purified SuCas12a2 over a HiLoad^TM^ 16/600 Superdex^TM^ 200 pg column. (B). SDS-PAGE of the recombinant SuCas12a2 with Coomassie brilliant blue staining. M stands for marker; lane 1 is the sample purified by Ni-NTA; lanes 2, 3, and 4 are the samples after molecular exclusion chromatography, corresponding to aggregates, monomers, and the last peak in A, respectively. (C). Schematic of non-target cleavage of pUC19 plasmid by activated SuCas12a2. (D). Concentration gradients and time-course analyses of RNA-triggered *trans* cleavage of non-target pUC19 plasmid. DNA was visualized and observed using ethidium bromide and 0.8% nucleic acid gel.

### 3.2 Construction of a diagnostic platform coupling PCR with SuCas12a2 and efficient detection of *Eh*Prx

*Eh*Prx was used to distinguish *E. histolytica* and *Entamoeba dispar* (*E. dispar*). Therefore, *Eh*Prx was used to test the feasibility of the assay. The extracted *Entamoeba* DNA was amplified using PCR primers tagged with a T7 promoter sequence. PCR products were detected using the CRISPR-Cas12a2 system. Fluorescence was generated after the Cas12a2 was activated and cleaved, which converts a nucleic acid signal that is difficult to detect into an easily detectable fluorescent signal (Figure 2a). To validate the specificity of this approach, we performed an experiment using *Eh*Prx gene-specific primers for *E. histolytica*, *E. dispar*, *Entamoeba nuttalli*, *Entamoeba moshkovskii*, and *Giardia* genomic DNA, and the products were purified. Subsequently, we introduced the clean-up product into the SuCas12a2 reaction mixture. Fluorescence was produced only in the presence of *E. histolytica* DNA, whereas other pathogens, even those belonging to other *Entamoeba* species, did not (Figure 2b, 2c). Importantly, the amplicons of *E. histolytica* and *E. dispar* were identical, although the PFS of *E. histolytica* differs by one base from that of *E. dispar*. This result indicated that the diagnostic method was highly specific, which was attributed to the precise recognition of PFS by SuCas12a2.

**Figure 2.**
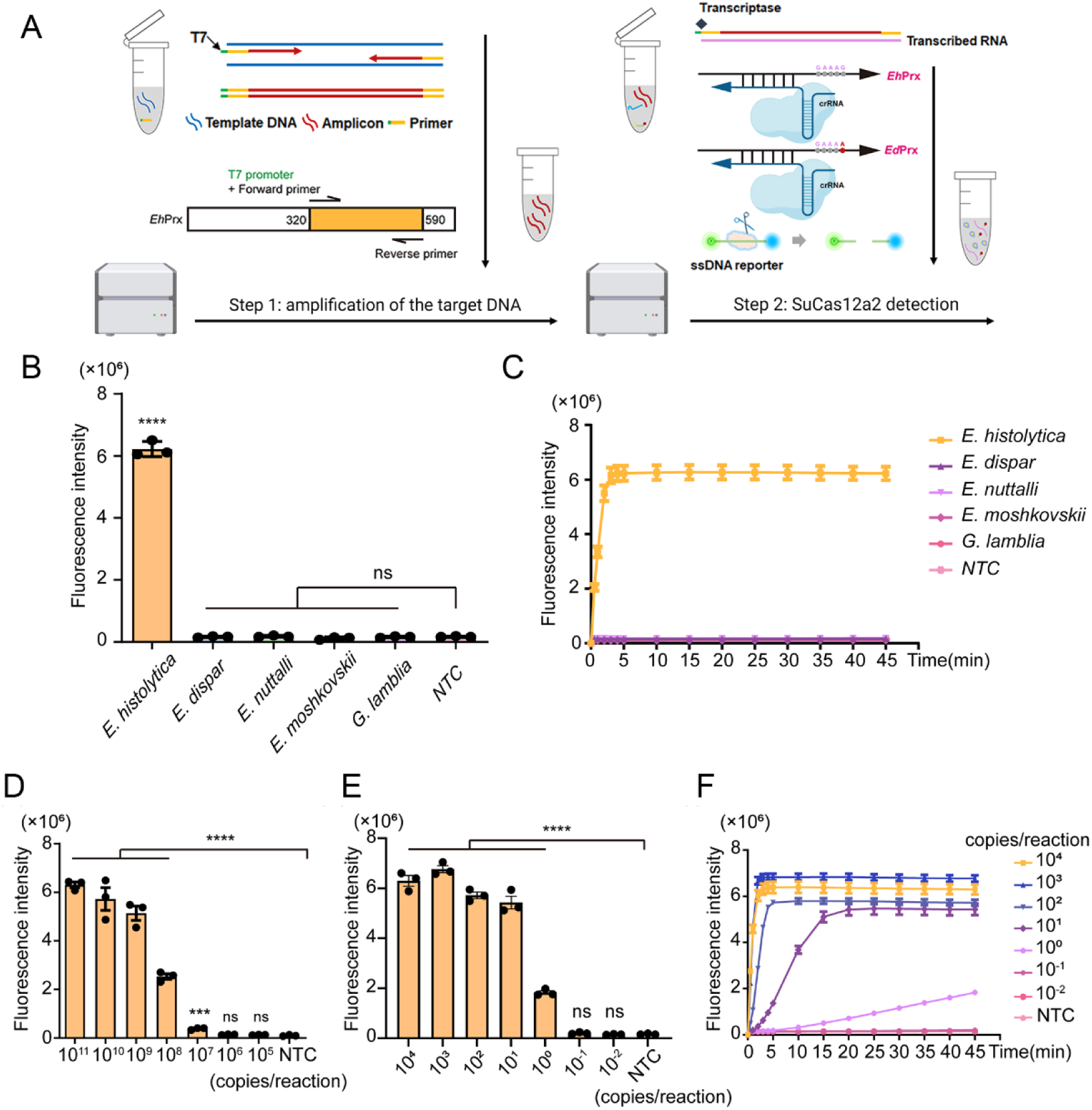
Evaluate the performance of PCR-SuCas12a2 assay using *E. histolytica* (A). Schematic of CRISPR/Cas12a2-based platforms. Target DNA fragments are amplified by PCR. The PCR products were purified and transferred into a new tube for SuCas12a2 detection, where the RNA is synthesized by amplicons harboring the T7 promoter sequence under the function of T7 RNA polymerase. SuCas12a2, crRNA, and specific RNA form a ternary complex, and fluoresces are generated when the RNA reporter cleaves. A single base change can distinguish the *E. histolytic* and *E. dispar* by PCR-SuCas12a2 assay. Assessment of the specificity of the PCR-SuCas12a2 assay through the detection of various amoeba and giardia species, evaluated via (B) end-point fluorescence intensity and (C) a 45-minute kinetic analysis. (D). Purified DNA of across various orders of magnitude was utilized to determine the minimum number of copies necessary for initiating the biological activity of SuCas12a2 nuclease. (E). End-point fluorescence intensity signal readout, along with a 45-minute kinetic analysis (F), was employed to evaluate the sensitivity of the PCR-SuCas12a2 assay by testing purified PCR production at different orders of magnitude. Data are represented as mean ± SEM of three technical replicates. \*\*\**p <* 0.001, \*\*\*\**p* < 0.0001 by *t* test, ns = not significant.

To evaluate the sensitivity of this method, we performed a minimal DNA copy number assay to detect SuCas12a2 activation. Various dilutions of *E. histolytica* PCR products were added to the SuCas12a2 assay environment, and each reaction was increased by an order of magnitude from 10^5^ to 10^11^ copies. The results showed that the lower limit of the minimum of SuCas12a2 for activation was 10^7^ copies (*p* = 0.0006), which was statistically significant compared to the next order of magnitude or the negative control; however, the terminal fluorescence value of 10^7^ copies was approximately one-seventh of the high copy group (Figure 2d). Therefore, the optimal DNA content for Cas12a2 activation is 10^8^ copies. Next, we quantified the substrate DNA concentrations ranging from 10^-2^ to 10^4^ for PCR to explore the LoD of this method. We found strong fluorescence in one copy (*p* < 0.0001); however, when we decreased the copy number to 0.1, the signal was reduced to levels not statistically different from the negative control, meaning this is beyond the LoD of this method (Figure 2e). In addition, a high copy number of *E. histolytica* leads to a sharp reduction in the time of arrival at the fluorescence value plateau, and 10^4^ copies can quickly reach this level in approximately 3 min (Figure 2f). Taken together, our PCR-SuCas12a2 diagnostic tool enables rapid and robust detection of *E. histolytica*, and the correlation of the fluorescent signal with DNA substrates indicates that our platform is an efficient tool for quantitative pathogen detection.

### 3.3 PCR-SuCas12a2 diagnostic platform for the detection of *E. histolytica* DNA in clinical samples

To determine the application of this novel diagnostic tool in patient samples, we evaluated the capability of our platform to detect *E. histolytica* nucleic acids extracted from feces or liver tissues of five patients who tested amoeba IgG antibody positive for *E. histolytica* with an IFA (Figure 3a). The fluorescence values of the two samples quickly reached a plateau within 10 min, which was comparable to the fluorescence signal with prolonged SuCas12a2 detection (Figure 3b). Although the fluorescence values of the remaining samples increased slowly, they were still considered positive, and there was a statistically significant difference between the samples and the negative control (*p* < 0.0001) (Figure 3c). Furthermore, we analyzed the fluorescence values between all samples and the negative control at 20 and 25 min and found that there were statistically significant differences at 25 min, even though the fluorescence signal continued to increase with the extension of SuCas12a2 detection (Figure 3d). In addition, samples with low fluorescence intensity in the IFA showed a relatively flat slope in the SuCas12a2 reaction fluorescence curve. Our assay showed a 100% positive detection rate in concordance with results obtained with conventional detection methods, suggesting its potential application in a clinical setting.

**Figure 3.**
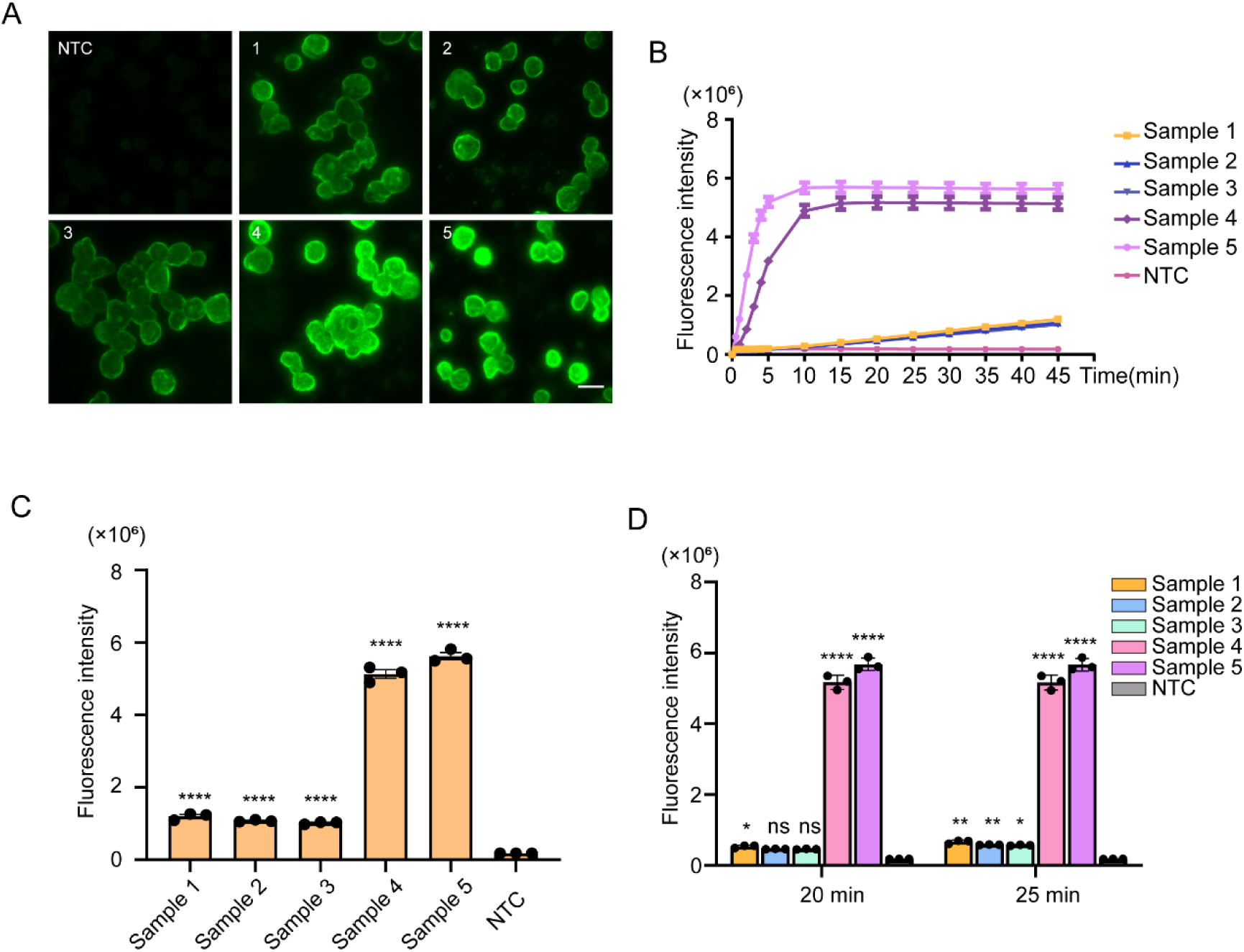
PCR-SuCas12a2 assay for detection of *E. histolytica* in clinical samples. A total of five samples were used to assess the clinical capacity of the PCR-SuCas12a2 assay. Fluorescence intensity at end-point (A) and 45-minute kinetic analysis (B) of five clinical samples were determined by PCR-SuCas12a2 assay. (C). IFA was used to determine five clinical samples, and the serum of healthy people was used as a negative control. (D). The fluorescence intensity of five clinical samples was detected by PCR-SuCas12a2 assay at 20 and 25 minutes, respectively. Data are represented as mean ± SEM of three technical replicates. \**p <* 0.05, *\*\*p <* 0.01, \*\*\*\**p* < 0.0001 by ordinary one-way ANOVA, ns = not significant; bar = 50 μm.

### 3.4 PCR-SuCas12a2 diagnostic platform optimization and practical application

Our platform, coupled with conventional PCR with SuCas12a2, reliably and robustly detected *E. histolytica* in clinical samples. To accelerate the detection process, we simplified our PCR-SuCas12a2 assay and attempted to develop a one-pot detection platform by introducing an RPA to replace the PCR. RPA was performed, and the product was directly added to the SuCas12a2 reaction for detection. Consistent with our expectations, the fluorescence template concentration for the sensitivity analysis, the results showed that each reaction harboring 10^5^ and 10^4^ copies could detect a strong fluorescence signal (Figure 4a), and the reaction endpoint was reached in 5 and 20 min (Figure 4b), respectively. The fluorescence signal values of 10^3^ and 10^2^ copies decreased significantly, and 10^1^ copies approached those of the negative control (Figure 4a). Thus, the LoD of the RPA-SuCas12a2 assay was 10^2^ copies per reaction, which was lower than the PCR-SuCas12a2 assay. Subsequently, we tested the specificity of the RPA-SuCas12a2 assay, which showed consistency with the PCR-SuCas12a2 method, with no cross-reaction with other amoebae and species (Figure 4c, 4d). Furthermore, all reaction components were added into one tube, utilizing 10^10^ copies as a template, to develop a one-pot detection platform, and a positive signal of *E. histolytica* was detected every 30 seconds for a total of 45 min at 37 ℃. The fluorescence signal appeared at approximately 20 min, and the final fluorescence value failed to match the two-pot assay, suggesting inferior sensitivity.

**Figure 4.**
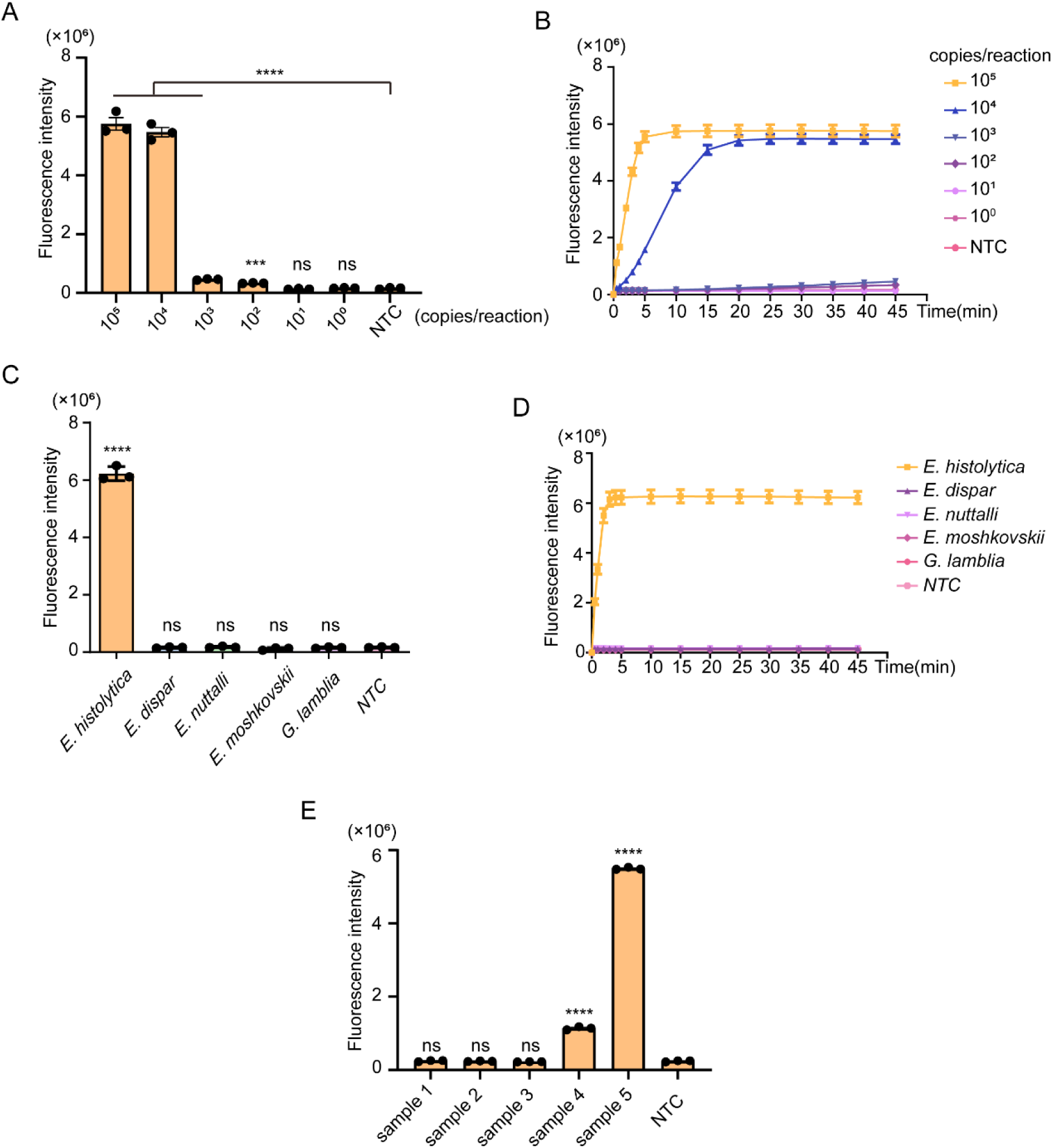
Evaluation the performance of the RPA-SuCas12a2 assay for the detection of *E. histolytica*. Sensitivity and specificity assessment of RPA-SuCas12a2 assay, (A & B) are the end-point fluorescence intensity and kinetic analysis for the LoD, respectively; (C & D) are end-point fluorescence signal and kinetic analysis for the specificity assessment among various amoeba, giardia species, and *E. histolytica*. (E). A total of five clinical samples were used and detected by RPA-SuCas12a2 assay. Data are represented as mean ± SEM of three technical replicates. \*\*\**p <* 0.001, \*\*\*\**p* < 0.0001 by ordinary one-way ANOVA, ns = not significant.

Next, we verified the feasibility of using RPA-SuCas12a2 to detect *E. histolytica* DNA in five clinical samples. The performance of the RPA-SuCas12a2 assay was inferior to PCR-SuCas12a2, with only one sample showing a strong fluorescence signal (Figure 4e), meaning its accuracy was significantly lower than that of the PCR-SuCas12a2 method. The RPA-SuCas12a2 assay is a powerful and robust nucleic acid detection system that can rapidly detect pathogens.

## 4. Discussion

In this study, we established a CRISPR-based detection platform that uses conventional nucleic acid amplification coupled with Cas12a2 as a robust tool for pathogen detection, using *E. histolytica* to assess the efficacy of this system. We constructed an expression plasmid via inserting the encoded *SuCas12a2* gene into the polyclonal site of the vector. The SuCas12a2 protein was successfully expressed and purified and showed efficient *trans*-cleavage activity, as indicated by cleavage of the pUC19 plasmid assay. The number and location of the encoded His-tagged genes had no apparent effect on protein function.

Nucleic acid detection platforms usually require high specificity and sensitivity [31]. In order to prevent false results, it is crucial to assess the performance of novel detection methods [32,33]. As a proof-of-concept, *E. histolytica* nucleic acids were detected to assess the specificity and sensitivity of the SuCas12a2-based system. Distinguishing *E. histolytica* and *E. dispar* has historically been difficult due to their high genomic similarity, as identified by primers existing in the *prx* gene [34], although the CRISPR/Cas12a2-based system can be implemented to distinguish these species. The PCR amplification products of *E. histolytica* and *E. dispar* were identical, with only a single-base difference in PFS sites, indicating that a precise match of the PFS is required for Cas12a2 activation. The results indicate that the CRISPR/Cas12a2-based diagnostic tool could reduce false-positive rates and may provide a potential tool in the detection of pathogen variants. SHERLOCK, based on Cas13 for ZIKV, and DETECTR, based on Cas12a for HPV, achieved attomolar sensitivity for pathogen detection [14,20]. The PCR-SuCas12a2 method is comparable, in which the LoD for *E. histolytica* detection is as low as one copy per reaction; this order of magnitude of copy number is sufficient for clinical diagnosis.

Cas12a2 recognizes RNA targets, unleashing non-specific dsDNA-, ssDNA-, and ssRNA-nuclease activities that are distinct from those of Cas12a, which is a dsDNA-targeting Cas nuclease [29,30]. Therefore, the CRISPR-Cas12a2 detection method demonstrates the advantage of rapid RNA detection, such as the diagnosis of various RNA viruses, which can be used in conjunction with RNA amplification technologies such as simultaneous amplification and testing for direct target RNA recognition using fluorescence quantification. SuCas12a2 is the nuclease of the RNA-targeting and can directly detect the RNA of the virus without transcription. While several tools have been developed based on Cas13, its technology is still in a developmental phase relative to the Cas12 family. There were several distinct differences between Cas12a2 and Cas13, although both are RNA targets. Cas12a2 can trim target RNA in *cis* conformations without degrading the hybridized target, thus enabling uninterrupted Cas12a2 activation [29]. This may be beneficial for the sustained activation of the Cas12a2 enzyme under low target RNA conditions and for boosting the fluorescence detection signal, which usually increases detection sensitivity. However, the release of non-specific dsDNA-, ssDNA-, and ssRNA-nucleases may make it more difficult to combine one-step detection, fusion amplification, and Cas12a2 detection in a single tube. Thus, RPA was used in our assay to replace PCR and amplify the target fragment. RPA-SuCas12a2 could also effectively detect target RNA, and this isothermal amplification technique could simplify the amplification process if an integrated chip was used to connect the two-step reactions in series. This may achieve the requirements of high-throughput detection.

## 5. Conclusions

A previous study developed nucleic acid scissors tools based on SuCas12a2, which could be used to diagnose variable mutation pathogens with accurate recognition performance and offer a promising alternative for pathogens surveillance. The sensitivity and specificity of the system based on Cas12a2 were excellent. Moreover, in cases where the tested sample exhibits a strong positive result or when the infection is in its early stages with a high viral load, these methods are capable of completing detection within 10 min. We are optimistic about further expansions based on the functions of the system, such as the practical application of RNA virus detection and the integration of rapid chip technology, to amplify the advantages of this enzyme for nucleic acid detection. Additionally, tools for the rapid detection of pathogens should be practical and scalable. Consequently, other versions based on the SuCas12a2 nuclease can be further developed, such as visual processing, immune chromatography technology, or chemiluminescence.

## Data Availability

All data produced in the present study are available upon reasonable request to the authors

## CRediT authorship contribution statement

X.C. conceived the idea and designed the experiments. H.Y. and M.F. performed the experiments. C.L., F.W., G.S., W.J. helped with the experiments. H.Y. drafted the manuscript. X.C., M.F., W.J. revised the manuscript. All authors read and approved the final manuscript.

## Data availability

Data will be made available on request.

